# Estimating the selection pressure of tumor growth on tumor tissue microbiomes

**DOI:** 10.1101/2024.03.17.24304406

**Authors:** Lianwei Li, Zhanshan (Sam) Ma

## Abstract

**Background:** The relationships between tumor and its microbiome are still puzzling, with possible bidirectional interactions. Tumor microbiomes may suppress or stimulate tumor growth on the one hand; on the other hand, tumor growth may exert selection pressure on its microbiomes. There is not any consensus on the mode and/or extension of the bidirectional interactions. The objective of this study is to estimate the selection pressure from the primary tumors on tumor microbiomes by comparing with the selection pressure from the solid normal tissues on their corresponding tissue microbiomes across 20+ cancer types.

**Methods:** We apply Sloan near neutral theory and big datasets of tumor tissue microbiomes from the TCGA (The Cancer Genome Atlas) databases to achieve the above objective. The near neutral theory model can determine the proportions of above-neutral, neutral and below-neutral species in microbial communities, corresponding with positive, neutral and negative selection pressures from host tissues. By comparing the proportions between the primary tumors and solid normal tissues, we can infer the selection pressure of tumor growth on tissue microbiomes.

**Results:** We find that approximately 65% of species in solid normal tissue microbiomes are neutral, and the proportion is only 40% in the primary tumor microbiomes. In contrast, the proportion of positively selected species exceeds 60% in the primary tumor microbiomes. Furthermore, simulations with neutral theory model reveal that most abundant species are mostly neutral, while non-neutral species are in the long tail of the species abundance distributions.

**Conclusions:** Tumor growth exerts strong positive selection on resident microbiomes, driving the abundances of certain species above the levels expected by the neutral process. Nevertheless, neutral species are still among the most abundant species, suggesting the necessity to pay close attention to the low-abundance or rare species because they are likely to play a critical role in oncogenesis.

## Introduction

The tumor tissue acquires functional abilities as it transitions from normal solid tissue, which are defined as hallmarks (Hanahan 2022). There are eight hallmarks, including sustaining growth signals, evading growth suppression, resisting cell death, enabling unlimited replication, inducing or accessing blood vessels, activating invasion and metastasis, reprogramming metabolism, and avoiding immune destruction. In their recent study, Hanahan (2022) also considered epigenetic reprogramming of non-mutagenic factors, as well as the diversity and heterogeneity of the microbiome, as new hallmarks. Researchers found distinct microbial signatures associated with different cancers (Poore et al. 2020). Machine learning models could accurately discriminate between cancer types and distinguish tumor vs normal samples based on the microbiome profiles alone. This held even after extensive validation checks and contamination removal. Analysis of blood samples also found predictive microbial signatures, including for early-stage cancers and cancers lacking detectable genomic alterations by commercial DNA tests (Poore et al. 2020, Zhao et al 2020).

Studies show that the microbiome plays an important part in tumor formation, cancer cell differentiation, and how fast cancer progresses. The microbiome interacts with and impacts other established characteristics of cancer, like tumor inflammation, evading immune system destruction, genetic instability, and resistance to cancer treatments (Lythgoe et al. 2022; Parida & Sharma 2021). Additionally, Certain pathogenic bacteria like *Helicobacter pylori, Chlamydia trachomatis*, and *Fusobacterium nucleatum* have been associated with increased risk of various cancers like stomach, cervical, and colorectal cancers respectively. Their genotoxins and other mechanisms can directly damage DNA and promote cancer. The confirmed microbial species that directly induce carcinogenesis are less than two dozen. Modulating the microbiome through diet, prebiotics, probiotics, fecal microbiota transplant etc. shows promise as an adjuvant cancer therapy approach. This can help maximize treatment benefits and minimize side effects. Certain bacteria like Salmonella, Clostridium and Bifidobacterium are being explored as targeted cancer therapies, either to directly kill tumors or deliver drugs specifically to the tumor. (Poore et al. 2020, Parida & Sharma 2021). The researchers also found that removing the intratumor bacteria significantly reduced lung metastasis without affecting primary tumor growth (Fu et al 2022).

Current research on the tumor tissue microbiome focuses on its diversity, heterogeneity, and interaction with cancer cells and the immune system. However, there is a lack of ecological research investigating the mechanisms that maintain diversity in the tumor tissue microbiome. It is unclear whether definitive or stochastic factors play a more important role in microbiome community assembly within tumors. The classic niche theory assumes that each species has its own niche. However, in some communities, there may not be enough niches to accommodate all species. To address these issues, several models for detecting the neutral process were proposed which focus on the stochastic birth, death and immigration in shaping community structure since Stephen Hubbell published his book about neutral theory (Hubbell 2001). Consequently, these neutral models were used to detect not only macro-ecology but also microbial ecology, such as the human microbiomes, hot spring microbiomes, milk microbiomes, lung microbiomes, and animal gut microbiomes, etc. (Venkataraman et al 2015; Li & Ma, 2016, 2019; Li et al., 2021; Ma, 2021).

This paper employs a neutral community model to evaluate whether microbial communities conform to the neutral process assembly. The research comprises two schemes: Scheme I compares solid tissue normal microbiomes and primary tissue microbiomes to detect differences in shaping community structure, while Scheme II simulates the transformation of microbiome communities from solid tissue normal microbiomes to primary tissue microbiomes. Investigates the role of deterministic and stochastic factors in the transformation of the microbiome community during tumor development by replacing spatial differential with temporal transformation.

## Methods and Datasets

### Cancer tissue microbiomes datasets

The original datasets we used in this research were published by Poore et al (2020), which re-examined whole-genome and whole-transcriptome sequencing studies in The Cancer Genome Atlas of 33 types of cancer. Details of how these samples were processed are described in Poore et al (2020). The raw reads were aligned to known human reference genomes, and any unaligned reads were mapped to bacterial, archaeal, and viral reference genomes using Kraken algorithm. This research aimed to investigate the role of deterministic and stochastic factors in the transformation process of the microbiome from solid tissue normal to primary tumor. The datasets must include both solid tissue normal and primary tumor samples in one type of cancer. So we selected samples from whole-transcriptome datasets covering 11 types of cancer, including 641 solid tissue normal samples and 6660 primary tumor samples. Each sample contained abundance of bacteria, archaea and virus. The samples contained an abundance of bacteria, archaea, and viruses. We conducted separate analyses for each taxon, as well as a combined analysis of the entire community.

### A Neutral Community Model for Microbial Community

The computational process and program for fitting neutral community model used in this research was from Venkataraman et al (2015) and Sloan (2006, 2007). The neutral model was published by Sloan (2006), they built a new tailored neutral model to test whether stochastic births, deaths and immigration have a key role in shaping the structure of prokaryotes based on assumptions of Hubbell (2001). The neutral community model can handle large populations and communities, which are common characteristics of microbiome communities. The aim of this model is to assess whether microbial communities fit with neutral community assembly. The following is a brief description on it.

Assuming there is a saturated community with a total of *N*_*T*_ individuals. Any individual who dies or leaves the community must be replaced by an immigrant individual from source community with probability *m*, or by an offspring of a member of the local community with a probability 1-*m*. The community undergoes transformation through the cycle of immigration, birth and death. For species *i*, with an initial abundance of *N*_*i*_, it may increase by one individual, remain unchanged, or decrease by one individual in a cycle of immigration, birth, and death with a certain probability:

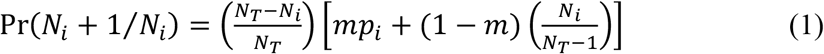

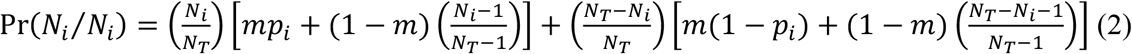

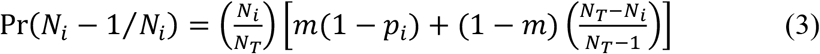

where *p*_*i*_ is the relative abundance of species *i* in the source community. For all species, the probability density function follows a beta distribution,

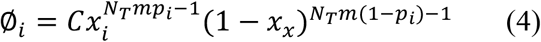

where, 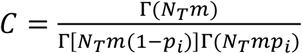, *x*_*i*_ is relative abundance of species in this local community.

The neutral community model allows us to determine whether each species is neutral or under selective. The process for testing the neutral community model as follows:

1. Compute *p*_*i*_ and *x*_*i*_ by fitting a beta distribution and estimating *m*.
2. Calculate the theoretical occurrence frequency of species *i* across all local community samples using *m* and the beta distribution.
3. Determine whether the observed *x*_*i*_ of species *i* falls within its 95% theoretical interval predicted by the neutral community model, and compile a list of neutral, below neutral, and above neutral species.

This model requires a source community and a destination community (i.e. a local community). Previous research has treated different spatial communities as both source and destination communities (such as mouse sites to lung microbial community, Venkataraman et al 2015). This study replaces spatial differential with temporal transformation and treats the solid tissue normal microbiome as the source community and the primary tumor microbiome as the destination community (local community) to investigates the role of deterministic and stochastic factors in the microbiome community transformation during tumor development. We preform the follow two schemes for this research.

Scheme I: Designate SNTM as both the source and destination communities simultaneously to detect which species in the SNTM assemble through neutral processes and which are subject to selective forces. The same should be done for PTM. Then compared the relative importance of neutral process and selective forces in SNTM and PTM.

Scheme II: To simulate the microbiome community transformation from SNTM to PTM, designate the SNTM samples as the source community and the PTM samples as the destination community. The above neutral species are positive selective species in PTM, and below neutral species might be the negative species in PTM. Display the composition of the PTM, find out which species are inherited from the SNTM with neutral drift and which species are under positive or negative selection. This study replaces spatial differential with temporal transformation to investigates the role of deterministic and stochastic factors in the microbiome community transformation during tumor development.

## Results

### Comparison of Mechanisms Underlying SNTM and PTM Community Assembly

We applied a neutral community model to compare the mechanisms shaping solid normal tissue microbiome (SNTM) and primary tumor microbiome (PTM) community assembly (Scheme I). Neutral models were fit separately to SNTM and PTM samples designated simultaneously as source and local communities (Tables S1-S2). Microbial communities comprised of archaea, bacteria, viruses, or all taxa were analyzed independently. For the results, all communities were split into three part: below neutral species, neutral species, and above neutral species. The below neutral species undergo negative selection, while the above neutral species undergo positive selection. The neutral species, on the other hand, are consistent with a neutral process.

In SNTM, the mean percentages of below-neutral, neutral, and above-neutral species were 7.99%, 69.3%, and 22.6%, respectively (Table S1). In contrast, these percentages in PTM were 17.7%, 38.4%, and 43.9% (Table S2). The percentage of below-neutral and above-neutral species was significantly higher in PTM, while the percentage of neutral species was significantly higher in SNTM (Table 1, Figure 1).

**Table 1.** The *p*-value of Wilcoxon test for percentage of below neutral, neutral and above neutral species between SNTM and PTM cohorts (Scheme I)

| Groups | Microbial Taxa | ≠ | > | < |
| --- | --- | --- | --- | --- |
| Below Neutral Species | Archaea | <0.001 | 1.000 | <0.001 |
|  | Bacteria | 0.001 | 1.000 | <0.001 |
|  | Viruses | <0.001 | 1.000 | <0.001 |
|  | Whole | <0.001 | 1.000 | <0.001 |
| Neutral Species | Archaea | <0.001 | 0.000 | 1.000 |
|  | Bacteria | <0.001 | 0.000 | 1.000 |
|  | Viruses | <0.001 | 0.000 | 1.000 |
|  | Whole | <0.001 | 0.000 | 1.000 |
| Above Neutral Species | Archaea | <0.001 | 1.000 | <0.001 |
|  | Bacteria | <0.001 | 1.000 | <0.001 |
|  | Viruses | <0.001 | 1.000 | <0.001 |
|  | Whole | <0.001 | 1.000 | <0.001 |

**Table 2.** The minimum, median, mean and maximum of relative abundance for below neutral, neutral and above neutral species.

| Cancer Types | Groups | Minimum | Median | Mean | Maximum |
| --- | --- | --- | --- | --- | --- |
| Breast Invasive Carcinoma | Below Neutral | 2.45E-09 | 1.26E-05 | 8.00E-05 | 1.15E-02 |
|  | Neutral | 6.09E-09 | 6.39E-05 | 2.84E-03 | 1.61E-01 |
|  | Above Neutral | 3.58E-08 | 1.39E-06 | 5.69E-06 | 1.59E-04 |
| Colon Adenocarcinoma | Below Neutral | 2.06E-08 | 2.89E-05 | 8.12E-05 | 1.32E-03 |
|  | Neutral | 4.13E-08 | 1.97E-04 | 4.35E-03 | 1.54E-01 |
|  | Above Neutral | 5.71E-08 | 2.21E-06 | 6.06E-06 | 6.10E-04 |
| Head and Neck Squamous Cell Carcinoma | Below Neutral | 1.01E-08 | 5.67E-06 | 8.06E-05 | 1.44E-02 |
|  | Neutral | 1.68E-08 | 4.32E-06 | 2.16E-03 | 1.41E-01 |
|  | Above Neutral | 6.91E-08 | 2.06E-06 | 1.08E-05 | 4.15E-04 |
| Kidney Renal Clear Cell Carcinoma | Below Neutral | 1.87E-08 | 1.51E-05 | 4.15E-05 | 1.55E-03 |
|  | Neutral | 7.01E-09 | 1.25E-04 | 3.32E-03 | 1.41E-01 |
|  | Above Neutral | 2.71E-08 | 9.26E-07 | 3.08E-06 | 9.13E-05 |
| Kidney Renal Papillary Cell Carcinoma | Below Neutral | 4.84E-09 | 7.65E-06 | 2.95E-05 | 9.59E-04 |
|  | Neutral | 8.69E-09 | 3.62E-06 | 1.82E-03 | 1.23E-01 |
|  | Above Neutral | 7.02E-08 | 1.32E-06 | 3.94E-06 | 1.03E-04 |
| Liver Hepatocellular Carcinoma | Below Neutral | 7.94E-09 | 4.21E-06 | 1.29E-04 | 3.42E-02 |
|  | Neutral | 7.11E-09 | 7.68E-07 | 1.59E-03 | 1.52E-01 |
|  | Above Neutral | 5.26E-08 | 1.39E-06 | 7.25E-06 | 2.45E-04 |
| Lung Adenocarcinoma | Below Neutral | 1.03E-08 | 2.51E-05 | 5.00E-05 | 1.82E-03 |
|  | Neutral | 6.17E-10 | 1.35E-04 | 3.10E-03 | 1.28E-01 |
|  | Above Neutral | 5.70E-08 | 1.17E-06 | 3.22E-06 | 1.44E-04 |
| Lung Squamous Cell Carcinoma | Below Neutral | 6.22E-09 | 1.22E-05 | 6.08E-05 | 7.46E-03 |
|  | Neutral | 8.08E-09 | 1.61E-05 | 2.12E-03 | 1.29E-01 |
|  | Above Neutral | 5.88E-08 | 1.64E-06 | 9.51E-06 | 1.18E-03 |
| Prostate Adenocarcinoma | Below Neutral | 6.81E-09 | 9.88E-06 | 3.04E-05 | 1.48E-03 |
|  | Neutral | 8.98E-09 | 7.68E-06 | 2.20E-03 | 1.47E-01 |
|  | Above Neutral | 5.82E-08 | 1.07E-06 | 3.43E-06 | 1.03E-04 |
| Stomach Adenocarcinoma | Below Neutral | 2.17E-10 | 1.84E-06 | 1.14E-05 | 1.46E-03 |
|  | Neutral | 6.46E-09 | 3.69E-05 | 2.21E-03 | 1.17E-01 |
|  | Above Neutral | 7.76E-09 | 3.84E-07 | 1.51E-06 | 1.53E-04 |
| Thyroid Carcinoma | Below Neutral | 7.08E-09 | 6.63E-06 | 2.84E-05 | 1.04E-03 |
|  | Neutral | 4.60E-09 | 6.62E-06 | 2.16E-03 | 1.15E-01 |
|  | Above Neutral | 5.44E-08 | 1.95E-06 | 9.53E-06 | 6.24E-04 |

**Figure 1.**
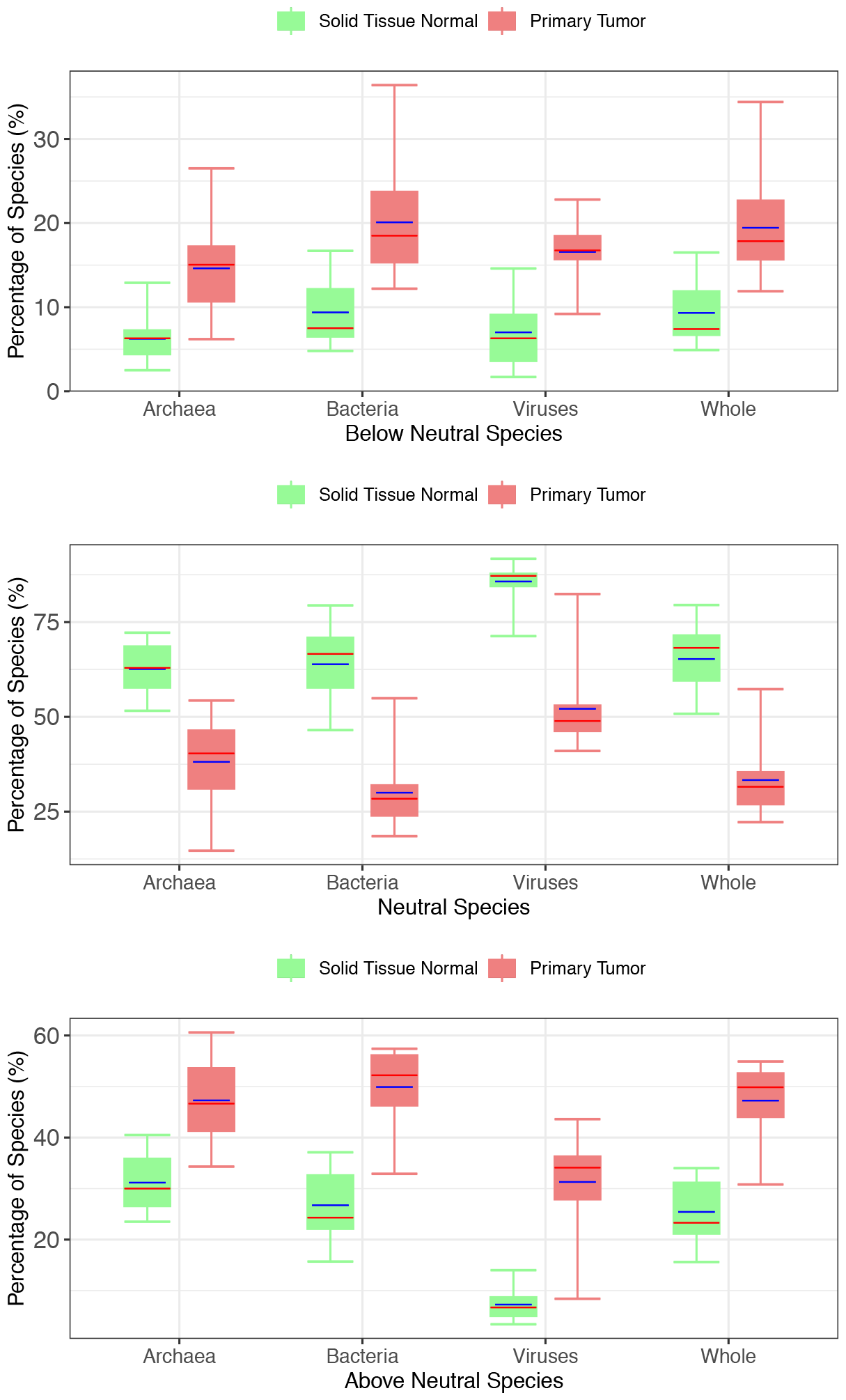
Box plots for percentages of below neutral, neutral and above neutral species for SNTM and PTM (Scheme I). The blue lines in box represent mean value across 11 types of cancer, and red lines are median. Standard bar charts showing the percentage of below neutral and above neutral species are significantly higher in the PTM than in the SNTM cohorts (*p*<0.001), while neutral species are significantly higher in the SNTM than in the PTM cohorts (*p*<0.001) (Table 1, Table S1, Table S2).

Across SNTM and PTM, viruses exhibited the highest percentage of neutral species compared to archaea and bacteria (Tables S1-S2). Even in PTM, the virus community had a higher neutral than above-neutral percentage. Archaea exhibit a higher *m* value, which represents the probability of a dead or leaving community individual being replaced by an immigrant from the source community, than bacteria and viruses.

### The Transformation from STNM to PTM Community

To characterize the mechanisms of microbial community construction or transformation during cancer development, we designed STNM as the source community and PTM as the destination community (Scheme II). The source and destination communities represent two temporal cancer development stages, rather than spatially distinct communities as previously assumed in the neutral community model. We want to use this neutral community model to distinguish which species are under neutral process and which are under negative or positive selection.

Table S3 displays the parameters and percentages of below-neutral, neutral, and above-neutral species for each microbial community. The percentage of above-neutral species is notably higher than neutral and below-neutral species (Figure 2), particularly for archaea and bacteria (more than 56%). The virus community exhibits an almost even distribution among the three categories (29.8%, 31.2%, and 39.0%,).

**Figure 2.**
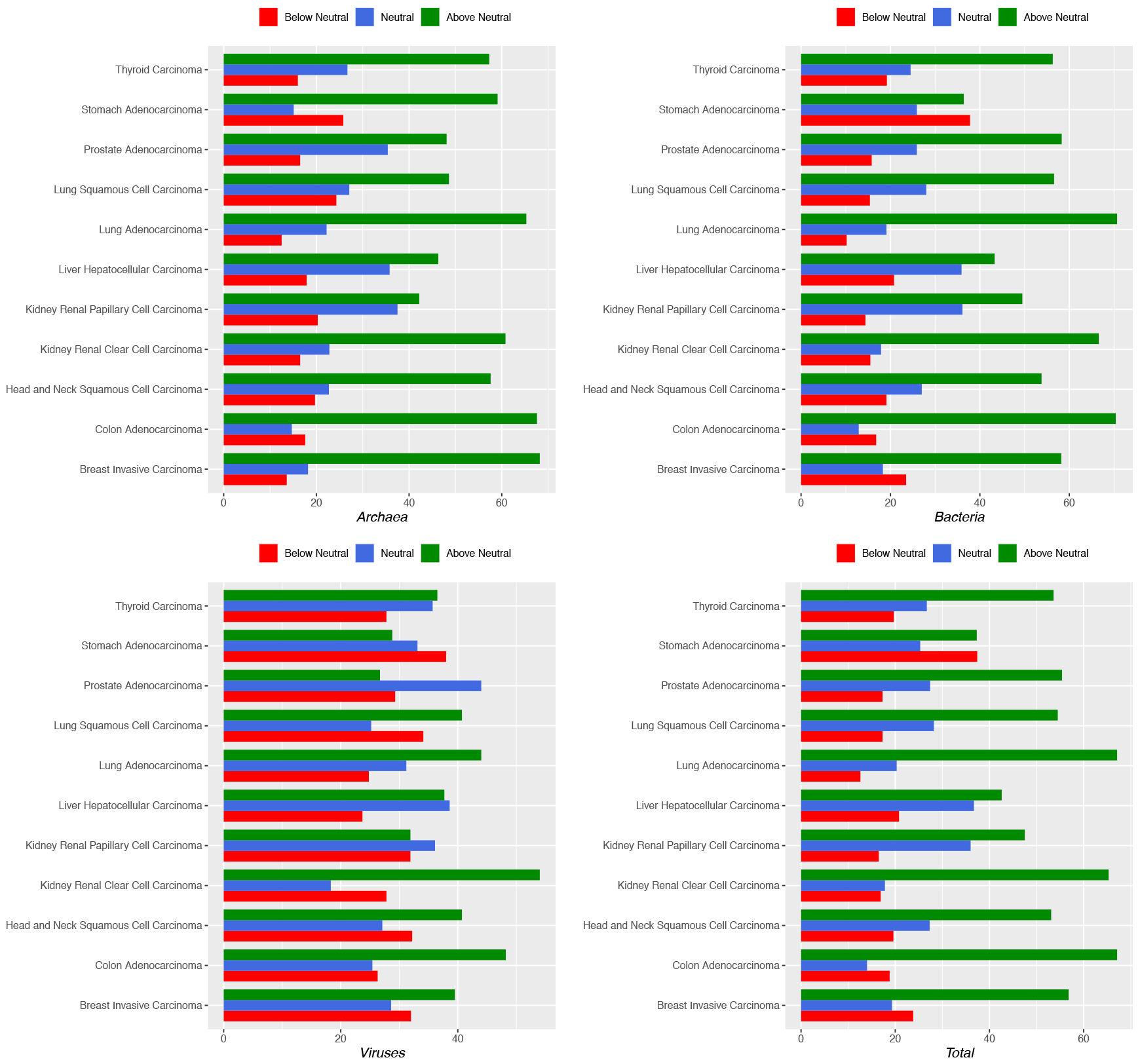
The percentage of below neutral species, neutral species, and above neutral species in PTM community. This neutral community model takes STNM community as source community and PTM as destination community.

Regarding the distribution of neutral, below-neutral, and above-neutral species in the PTM community, it can be concluded that almost all of the top relative abundance species are neutral species (as shown in Figure 3). The following are below-neutral species, and the most above-neutral species are located in the long tail.

**Figure 3.**
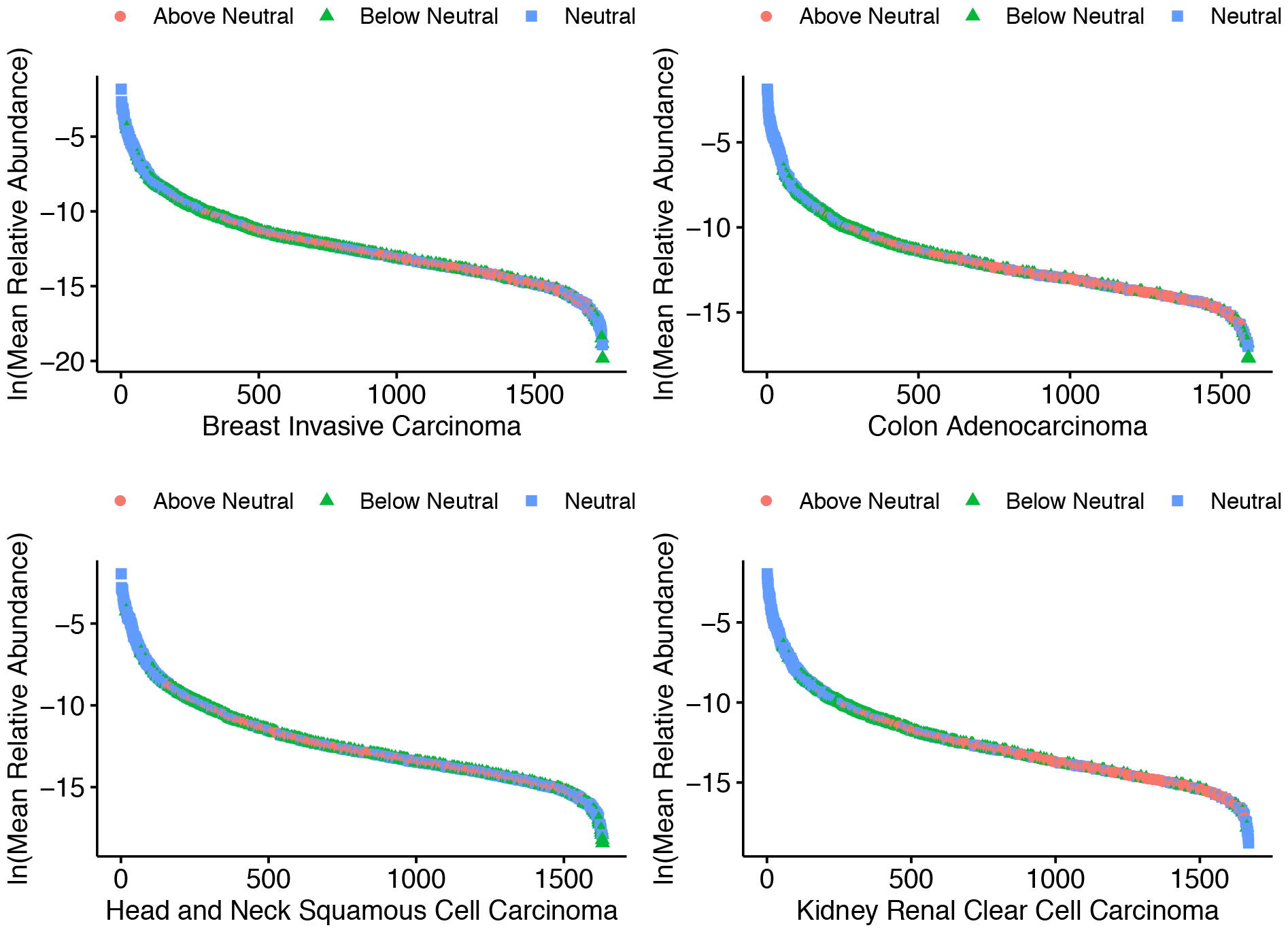
The species rank relative abundance of tumor tissue microbiomes. Neutral species are represented by blue points, below neutral species by green points, and above neutral species by red points. The y-axis represents relative abundance in nature logarithm of species; the x-axis represent species rank order in relative abundance.

## Data Availability

https://pubmed.ncbi.nlm.nih.gov/32214244/

